# Utilization of CoRDS Registry to Monitor Quality of Life in Patients with VCP Multisystem Proteinopathy

**DOI:** 10.1101/2024.08.18.24311141

**Authors:** Eiman Abdoalsadig, Merwa Hamid, Allison Peck, Leepakshi Johar, Virginia Kimonis

**Author notes:** Contributed equally to the work and should be considered co-first authors (Eiman Abdoalsadig, Merwa Hamid, Allison Peck). Correspondence: Virginia Kimonis, MD. MRCP Professor, Department of Pediatrics Division of Genetics and Genomic Medicine University of California-Irvine Lab and FEDEX: Hewitt Hall, Rm 2038, Health Sciences Rd., Irvine CA 92697.

## Abstract

**Background:** VCP disease, also known as multisystem proteinopathy (MSP1), is a rare, autosomal dominant, adult-onset, neuromuscular disease that is caused by variants in the valosin-containing protein (*VCP*) gene. VCP disease may exhibit one or more of the following primary features: Inclusion Body Myopathy (IBM), Paget’s disease of bone (PDB), Frontotemporal Dementia (IBMPFD), and Amyotrophic Lateral Sclerosis (ALS). Due to its progressive nature, death normally occurs in their sixties due to respiratory and cardiac failure. The purpose of this study is to utilize the Cure VCP Disease patient registry hosted by the Coordination of Rare Diseases at Sanford (CoRDS) to conduct a prospective natural history study.

**Methods:** Seventy-nine participants enrolled in the patient registry and answered demographic, *VCP* variant type, Patient-Reported Outcome Measures (PROMs), and Quality of Life (QOL) questionnaires over the course of three years. We additionally investigated if any sex differences existed and if genotype-phenotype correlations affected the rate of progression of the varying clinical manifestations.

**Results:** Overall, participants’ mobility declined significantly as the disease progressed. Participants reported a 0.6% decline in upper extremity function, 1.2% decline in lower extremity function, and 0.3% decline in cognitive function per year of age. Furthermore, participants reported a 1.6% decline in upper and lower extremity function and a 0.1% decline in cognitive function per year of disease duration. The highest PROMs correlations between overall health and lower extremity function, upper extremity function, fatigue, and the ability to perform vigorous activities. Genotype-phenotype correlations revealed no significant differences except for the absense of PDB in the *p*.*Arg159Cys* group.

**Conclusion:** The VCP CoRDS Registry was found to be a valuable tool for monitoring the QOL in patients with VCP disease and capturing patient perspectives for future clinical trials.

## Background

Valosin-containing protein (VCP) disease, otherwise known as inclusion body myopathy (IBM) associated with Paget disease of the bone (PDB) and frontotemporal dementia (FTD) (frequently used acronym, IBMPFD), or more recently, multi-system proteinopathy type 1 (MSP1), is a progressive, debilitating, predominantly autosomal dominant genetic condition, which can affect the muscle, bone, and/or brain. The condition was first reported in 2000 [1] as a new familial disorder of limb-girdle type of muscle weakness and skeletal disorganization due to PDB, which was later identified to be due to heterozygous missense variants in the *VCP* gene mapped to chromosomal location 9p211-p12 [2–4]. The *VCP (p97)* gene encodes for an abundant cytosolic AAA ATPase with multiple cellular functions. It is involved in the ubiquitin-proteasome system (UPS), the endoplasmic reticulum-associated protein degradation (ERAD), DNA endosomal sorting, cell cycle regulation, and autophagy [5–8]. To date, more than 80 missense variants associated with VCP disease have been identified [6]. At the cellular level, variants lead to the increased enzymatic activity of AAA ATPase reflecting a gain of function mechanism which dysregulates protein degradation through ERAD and/or UPS resulting in activation of the ER stress response [7] resulting in a global disturbance of protein folding [5].

Al-Obeidi et al. (2018), reported myopathy (typically IBM) in 90% of studied individuals with VCP disease, PDB in 42%, and FTD in 30% [8]. IBM manifests as progressive muscle weakness, difficulty in raising the arms and walking upstairs, and atrophy of the muscles of the pelvis and shoulder girdle. Histologically, IBM is characterized by rimmed vacuoles containing protein aggregates of TDP-43 [9]. PDB often consists of an increase in osteoclastic activity resulting in disorganized bone structure and bone weakness, resulting in fractures and localized arthritis [10]. *VCP* gene variants have also been reported to cause ALS in 10%, Parkinson’s disease in 5%, and cardiomyopathy in up to 27% of patients [8] [11–14]. FTD affects speech production and recall, comprehension, and personality [13–14]. Owing to the progressive nature of the disorder, death usually occurs prematurely in the 60s due to respiratory and cardiac failure [8].

Currently, while bisphosphonates can be effectively used to manage PDB, there are no treatments that can modify the progression of VCP disease-associated myopathy or FTD [6]. Medical management includes physical therapy, weight control, respiratory aids, and mechanical aids such as walkers or wheelchairs [6]. Patients are also recommended to receive regular surveillance to monitor cardiac, bone, lung, and neurologic function following diagnosis [6].

Even though VCP disease is now reported in several hundred patients across the globe, there are limited studies investigating how VCP disease affects the quality of life (QOL) of patients over time. We therefore planned to analyze patient reported data to ascertain the prevalence, age of onset of different manifestations, and track the progression of mobility, cognitive function, and pain levels in patients with VCP disease.

## Methods

Cure VCP Disease, Inc., a patient advocacy group established to unite patients and researchers to advance treatments and cures for VCP disease, collaborated with the Coordination of Rare Diseases at Sanford (CoRDS), a centralized international patient registry for rare diseases, to host a patient registry (VCP CoRDS Registry). The registry survey contains 143 unique questions related to demographics, diagnosis, age of onset, symptoms, disease progression, pain, and QOL. The registry participants are prompted to update survey responses annually. Thus, a longitudinal natural history study over the course of up to three years was established to explore the progression of QOL symptoms among VCP disease patients. Using data contained in the registry, we conducted the following analyses: 1) progression of functional abilities, 2) quality of life correlations, and 3) genotype-phenotype correlations. We compiled the genetic variant and phenotype responses from the VCP CoRDS Registry for each participant to conduct a genotype-phenotype comparison for the specific VCP disease variant. We used Excel and SPSS to conduct one sample t-tests with a 95% confidence interval for the statistical analysis in the genotype-phenotype comparison (IBM SPSS Statistics, Version 27).

For the progression of functional abilities analysis, we extracted from the VCP CoRDS registry, the responses from three surveys developed by the National Institute of Neurological Disorders and Stroke (NINDS): lower extremity function (Neuro-QOL Item Bank v 1.0 short form), upper extremity function (Neuro-QOL Item Bank v 1.0 short form), and cognition function (Neuro-QOL Item Bank v 2.0 short form) [15]. Each Neuro-QoL survey consists of eight questions developed to assess upper extremity function (such as their ability to turn a key in a lock, brush teeth, write with a pen or pencil, open and close a zipper), lower extremity function, and cognitive function, respectively. These patient-reported outcome measures (PROMS) have been previously proven to be valuable tools to increase understanding of disease symptoms and overall quality of life [16–17]. The 5-point Likert scale was applied in these responses to understand the extent to which participants can perform specific daily functions where a score of 1 is defined as the inability to perform a task while a score of 5 is the ability to complete a task without any difficulty. The total raw scores from the short forms were calculated for lower extremity function, upper extremity function, and cognitive function. To analyze pain in this population, we used the pain levels reported in the patient registry on a scale of 0-10 with 10 being the worst pain imaginable. Mild pain was defined as 0-3; moderate pain was 4-6; and severe pain was 7 or higher on the pain scale. We then compared pain levels and progression of pain between male and female participants by comparing their pain level with age as well as disease duration. We applied a linear regression model to observe the progression in mobility, cognitive function, and pain with age and disease duration.

Lastly, we analyzed the possible effect of disease progression, pain, and age on QOL. Participants reported their QOL symptoms by answering five questions in the QOL section of the VCP CoRDS Registry. We converted the participants’ responses using a 5-point Likert scale (where 1 represents poor QOL while 5 represents excellent QOL utilizing their latest responses), in which those scores were added to a bar chart comparing the latest responses of symptomatic and pre-symptomatic participants. Mobility and cognitive function were classified according to the total raw scores provided by the Neuro-QOL surveys. We calculated the Spearman correlation coefficients to evaluate if any correlations existed between each of the following nine metrics through a correlation matrix: age, disease progression (lower extremity, upper extremity, cognitive function, and pain), and QOL factors (vigorous activities, fatigue, general health, pain interference, and depression). We classified correlations between these factors as strong (± 0.7-1.0), moderate (± 0.4-0.6), and weak (± 0.0-0.3). We utilized the R program to generate a visually informative heat map, offering a comprehensive depiction and comparison of the various correlation coefficient magnitudes [R version 4.4.0 (Puppy Cup) prerelease versions].

## Results

Demographic information was collected from the VCP CoRDS Registry (Table 1). Seventy-nine participants [males (N=35, 44.3%), females (N=39, 49.4%), and five unreported sex (6.3%)] contributed to this study and approximately 89% of the participants reported themselves as ‘White’ with 4% ‘Asian’, and 6% ‘Mixed or Other’. The average age reported associated with diagnosis of muscle weakness, PDB, and dementia was 42.3 years, 47.3 years, and 54.8 years, respectively. Seventy-nine participants were registered from nine different countries (Figure 1). Most of the participants were from the United States (N=59, 71%), followed by the United Kingdom (N=5, 6%) and Australia (N=5, 6%). Four participants did not indicate their country of submission.

**Table 1.**
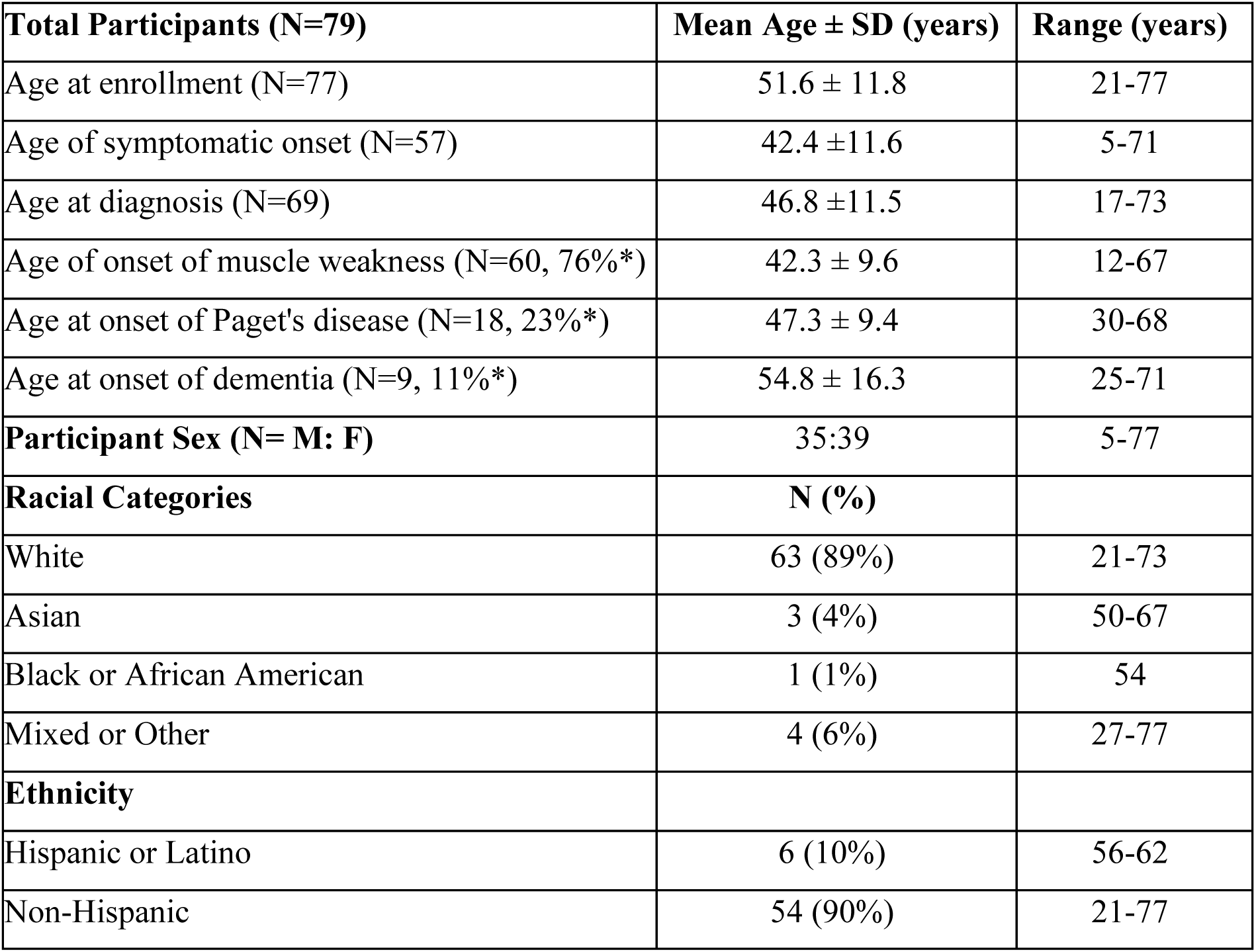
Demographic Data of Participants (* prevalence)

**Figure 1.**
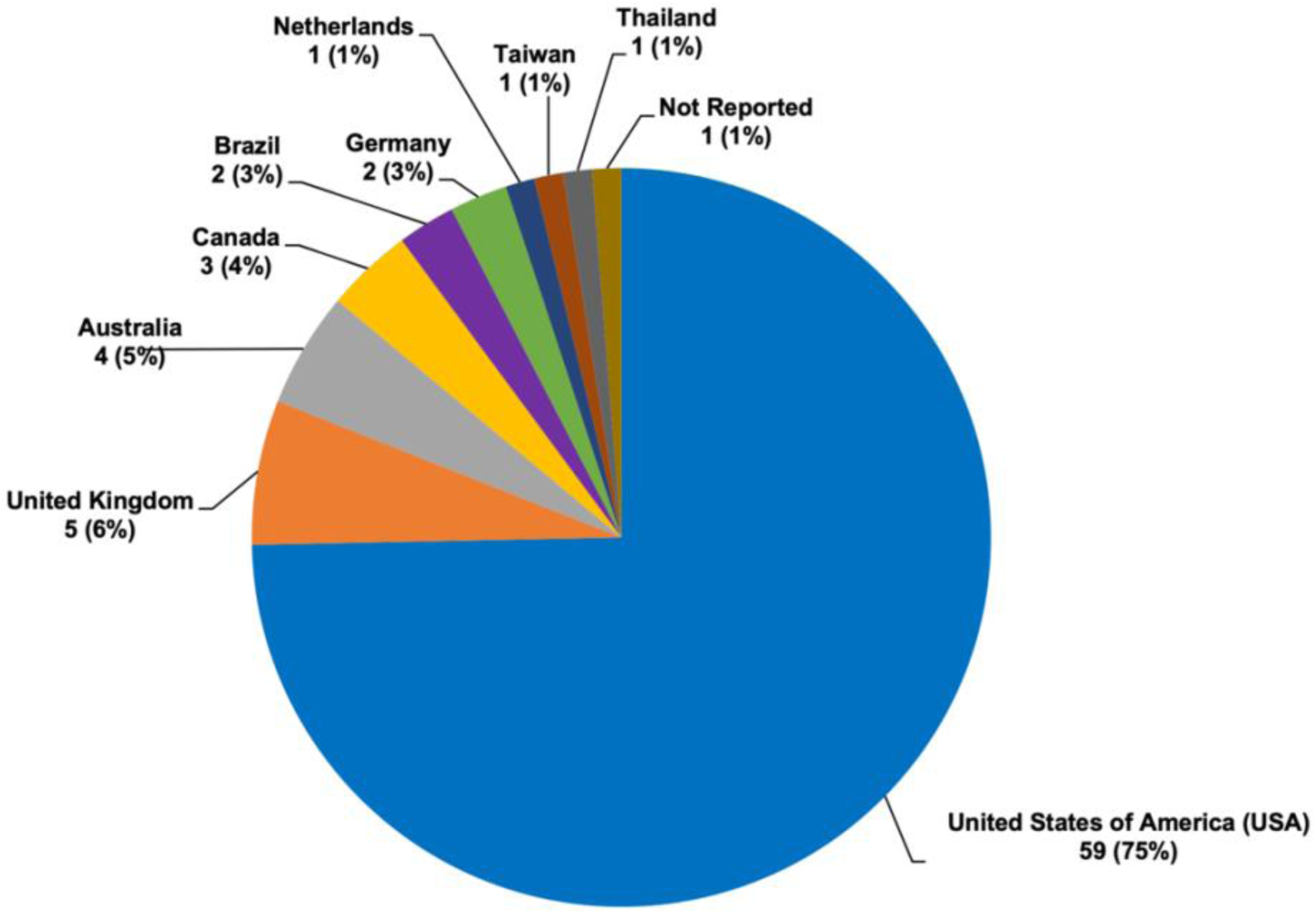
Demographic Data of Participants by Country of Residence. Seventy-nine participants from nine different countries participated in the study. Fifty-nine participants (75%) registered in the United States. One participant (1%) did not indicate country of submission.

Of the total number of participants registered, nine participants were pre-symptomatic (Figure S1). A pre-symptomatic participant is defined as someone who has a variant on the *VCP* gene but has neither been diagnosed with nor experienced a VCP-related clinical symptom. Sixty-six (84%) participants were symptomatic, meaning the participant either reported a clinical diagnosis or VCP-related clinical symptoms in the registry (Figure 2). Three participants indicated “other” under physical diagnosis, however, did not specify their comorbidities. The status of four participants is unknown. Out of the 62 symptomatic participants that reported a phenotype diagnosis, the most common clinical feature reported was myopathy (N=52, 83.9%); notably, of these patients, twenty participants (32.3%) reported isolated myopathy and thirteen participants (21.0%) reported both myopathy and PDB (Figure 2). The second most common physical diagnosis was PDB (N=18, 29.0%). In addition, nine participants reported FTD (14.5%), six participants reported peripheral neuropathy (9.7%), nine participants reported cataracts (14.5%), four participants reported cardiomyopathy (6.5%), three participants reported ALS (4.8%), and one participant reported Parkinson’s disease (1.6%). Sixty-seven participants reported their mean age of diagnosis for VCP disease as 49 years (range 23-73). Of the symptomatic group, twelve participants (17.9%) were diagnosed between the ages of 31-40 years, twenty-one (31.3%) were diagnosed between the ages of 41 years and 50 years, and twenty participants (30%) were diagnosed between the ages of 51 years and 60 years. A large majority of participants reported symptom-onset during adulthood; however, one participant reported having symptoms starting at the age of 12 years while another participant was pre-symptomatically diagnosed at the age of 17 years (Figure S2). Among the 56 participants who reported their age of first symptom onset, the most common age range was between 31-40 years (N=20, 36%), closely followed by 41-50 years (N=17, 30%). In this cohort, the average time from symptom onset to establishment of the diagnosis was 6.8 years with the longest duration of 29 years (Figure S2).

**Supplemental Figure 1.**
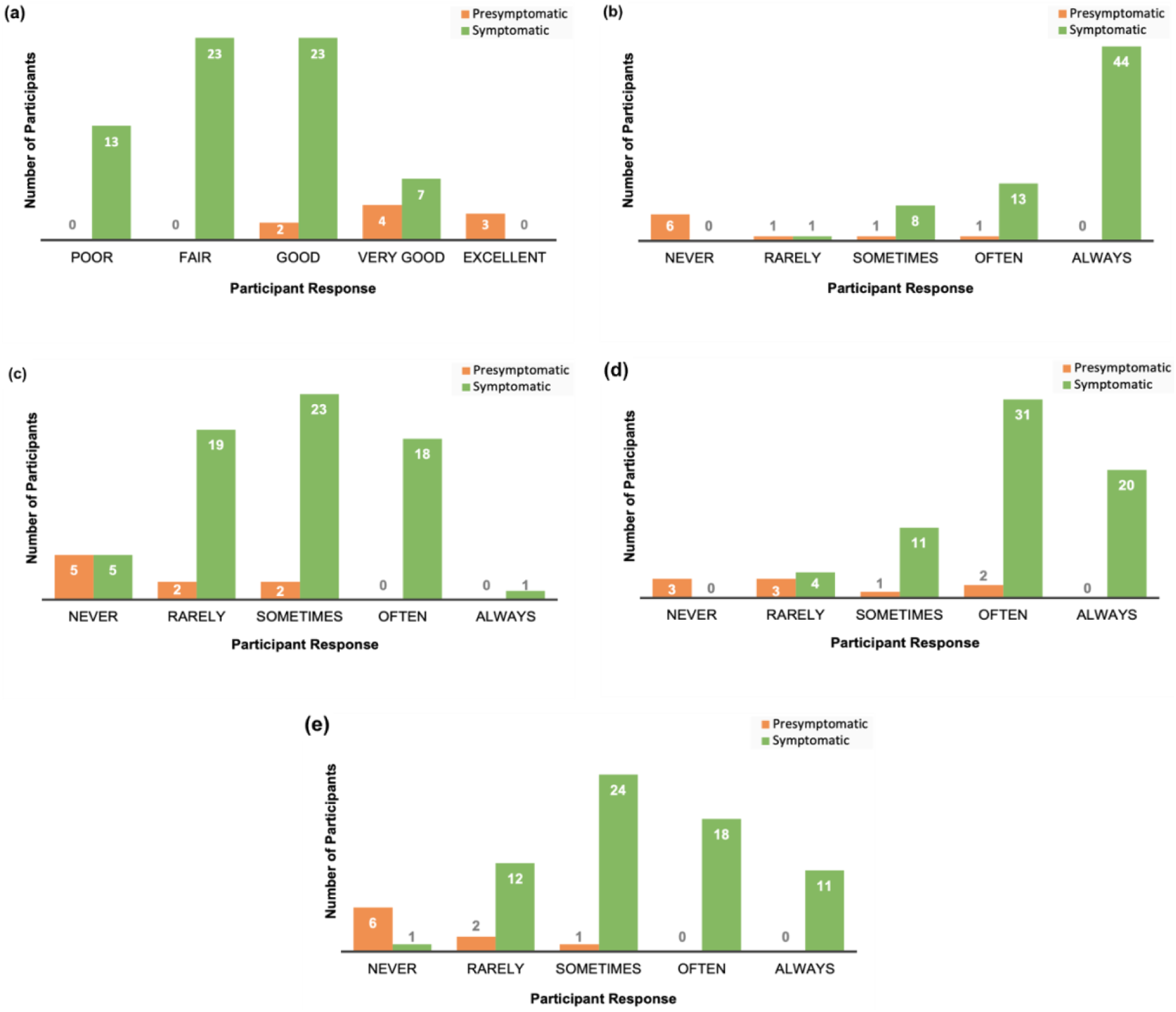
Quality of Life - A Comparison of Pre-Symptomatic vs. Symptomatic VCP Patients. A comparison between pre-symptomatic (orange, N=9) and symptomatic (green, N=66) participants when asked five QOL questions was conducted. **(a)** Participants’ responses when asked about their general health. **(b)** Participants’ responses when asked if their health limits them in doing vigorous activities. **(c)** Participants’ responses when asked if they feel depressed. **(d)** Participants’ responses when asked how often they feel tired. **(e)** Participants’ responses when asked how much pain interfered with their enjoyment of life.

**Figure 2.**
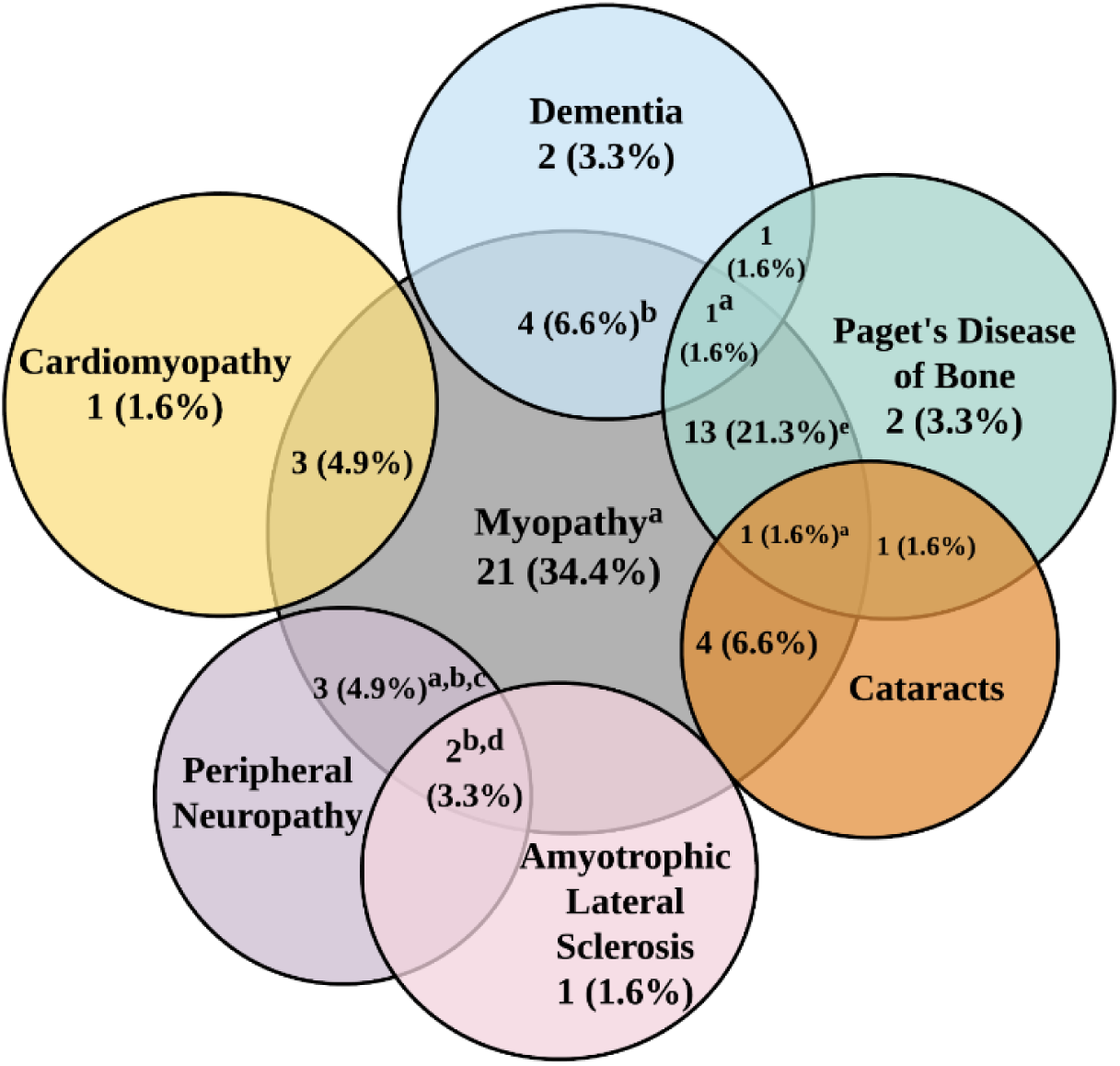
Venn Diagram of Physician-Confirmed Diagnoses in 66 Participants. Each circle represents a different physical diagnosis. Areas where the circles overlap are defined as the participant reporting having multiple diagnoses. One participant (not illustrated) who reported “other” was diagnosed with Parkinsonism and ataxia. The status of four participants (not displayed) is unknown. a: Indicated other in addition to provided physical diagnoses. b: One participant also reported cataracts. c: One participant also reported dementia. d: One participant also reported Parkinson’s disease. e: One participant also reported peripheral neuropathy.

**Supplemental Figure 2.**
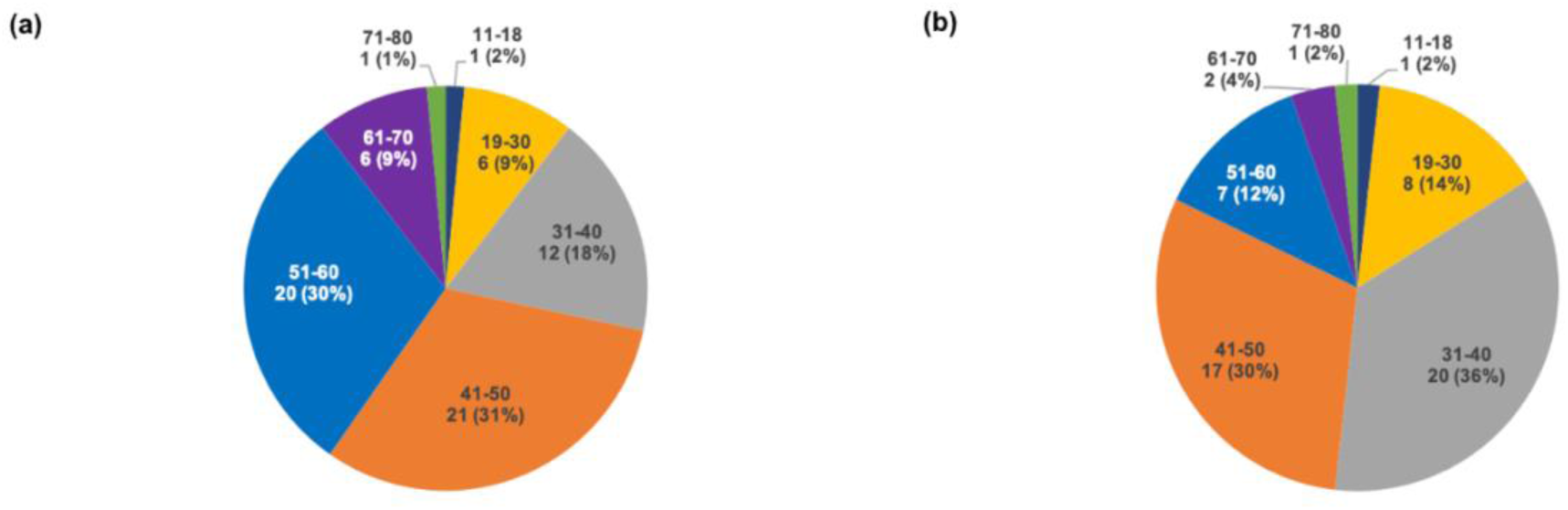
Demographics. **(a)** Age of diagnosis of participants with VCP disease (N=67). **(b)** Age of first symptom onset in VCP disease participants (N=56). Each slice represents a specific age range.

### Genotype-Phenotype Comparison

The genotype-phenotype analysis of this cohort revealed phenotypic diversity in VCP disease in relation to the variant group and the corresponding clinical features reported (Table 2, S1). Among the 49 participants (mean age 52.5 years) who reported their *VCP* variant, 17 (35%) reported the *p.Arg155His, c.464G>A,* variant, making this the most prevalent variant in this cohort (Table 2, S1). Not surprisingly, all the phenotypes were represented in this *p.Arg155His* cohort (myopathy, PDB, dementia, ALS, peripheral neuropathy, Parkinson’s disease, and cataracts); Parkinson’s disease was reported by only one participant in the R155H variant group in 5.6% (2%), ALS was only reported by two (4.1%) participants with *p.Arg155His* variant and was not reported in the other variant groups. The earliest reported age of onset was by a 27-year-old participant with myopathy and peripheral neuropathy with the *p.Arg155His, c.464G>A* variant (Table S1). The second most prevalent variant group with 8 participants was the *p.Arg155Cys, c.463C>T* group. Among all the variant groups, myopathy was identified in 36 participants [73.5%, 43.1 ± 8.8 years], making it the most common phenotype. The only possible notable genotype-phenotype correlations identified was the absence of PDB in the *p*.*Arg159Cys, c.475C>T* variant group.

**Table 2.** *VCP* gene variants in current cohort

| Variant group | N=49 (%) |
| --- | --- |
| p.Arg155His, c.464G>A | 18 (36.2%) |
| p.Arg155Cys, c.463C>T | 6 (17.0%) |
| p.Arg159His, c.476G>A | 5 (6.4%) |
| p.Arg93Cys, c.277C>T | 3 (8.5%) |
| p.Arg159Cys, c.475C>T | 4 (8.5%)<sup>b</sup> |
| p.Arg191Gln, c.572G>A | 3 (6.4%) |
| p.Leu198, c.Trp 593T>G | 2 (4.3%) |
| p.Gly97Glu, c.290G>A | 1 (2.1%) |
| p.Ile369Thr, c.1106T>C | 1 (2.1%) |
| p.Arg95Gly, c.283C>G | 1 (2.1%) |
| p.Asn387Thr, c.1160A>C | 1 (2.1%) |
| p.Gly128Ala, c.383G>C | 1 (2.1%) |
| p.Gly782Ala, c.2345G>C | 1 (2.1%) |

### Progression of Functional Abilities

#### Upper and Lower Extremity Function

We assessed the mobility of the sixty-seven participants who completed all the questions in the Neuro-QoL upper and lower extremity function surveys by analyzing the responses separately. Examples of PROM questions pertaining to upper extremity function include the ability to turn a key in a lock, brush teeth, write with a pen or pencil, open and close a zipper, etc. Affected participants showed an overall decline of 0.58% in upper extremity function with every 1-year increase of age (slope=-0.38, R^2^=0.23) (Figure 3a). Participants also displayed an overall decline of -1.57% in upper extremity function for every 1 year of disease progression (slope=-0.63, R^2^=0.32) (Figure 3b). Similar trends were seen in males and females with age (p=0.45) and disease duration (p=0.57).

**Figure 3.**
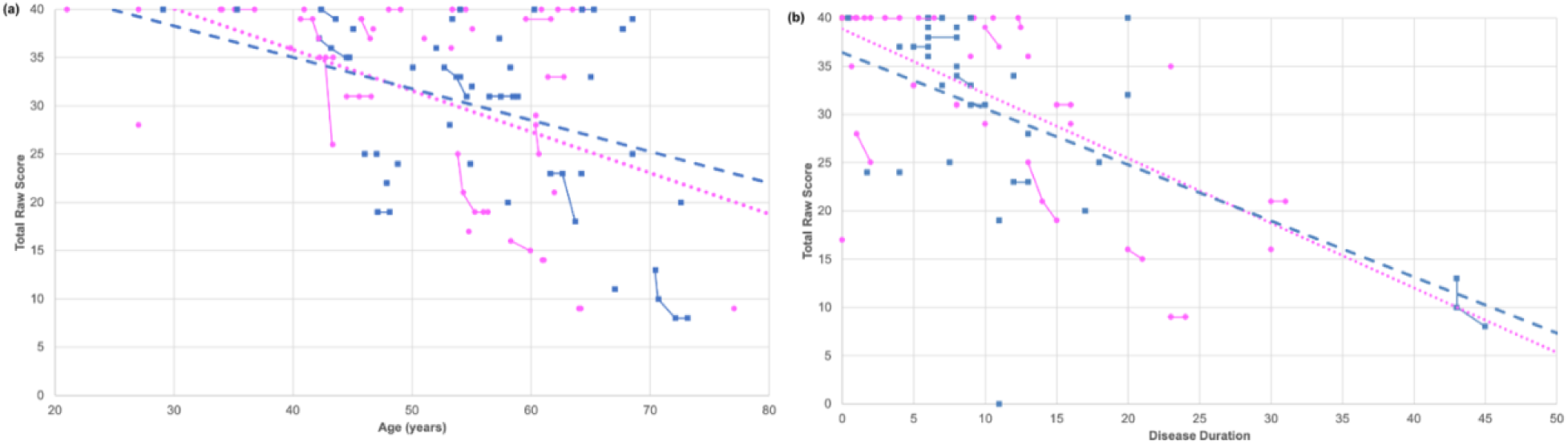
Upper Extremity Function Progression. A scatter plot was created to illustrate the correlations between **(a)** age and **(b)** disease duration relative to the total reported raw score for upper extremity function. The solid lines indicate progression within a singular participant over time for data with multiple submissions whereas the dashed lines indicate overall progression in males (blue) and females (pink) using the latest submission. Both male (blue squares) and female participants (pink circles) showed a rapid decline in their overall mobility with increasing age and also disease duration.

Both sexes exhibited similar trends in lower extremity function with increasing age with an overall decline of 1.19%, (Figure 4a). Participants reported an overall decline in lower extremity function as their disease progressed (slope=-0.64, R^2^=0.27) (Figure 4b). Females demonstrated a more rapid decline (slope=-0.66, R^2^=0.29) than males (slope=-0.59, R^2^=0.25). Participants also rated their upper extremity function by answering PROM questions such as their ability to run errands, get in and out of a car, step up and down curbs, and walk for at least 15 minutes.

**Figure 4.**
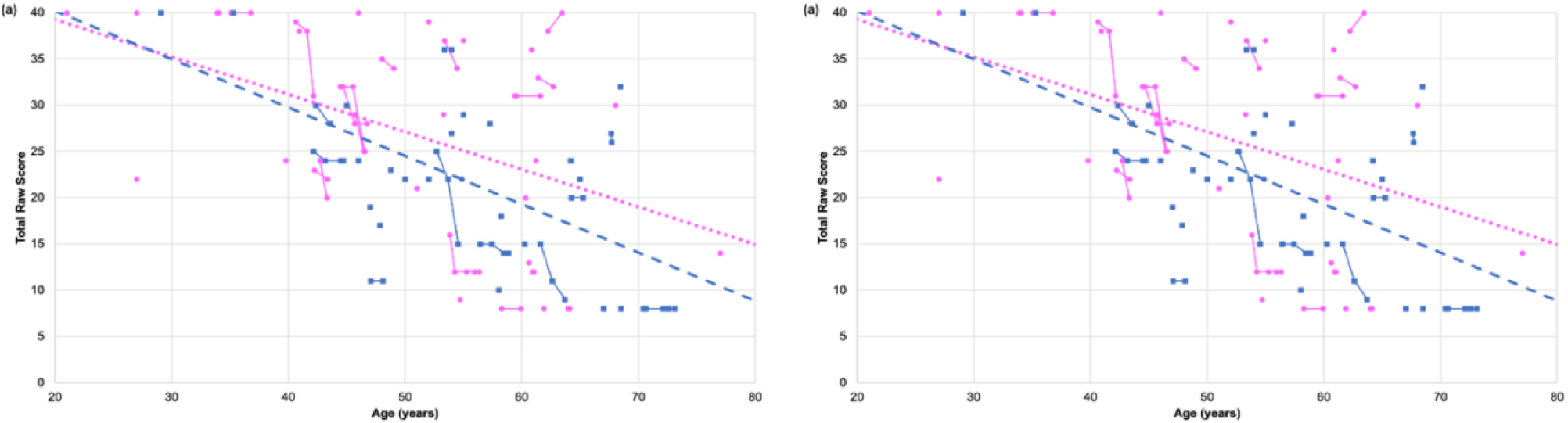
Lower Extremity Function Progression. A scatter plot was created to illustrate the correlations between **(a)** age and **(b)** disease duration relative to the total reported raw score for lower extremity function. The solid lines indicate progression within a singular participant over time for data with multiple submissions whereas the dashed lines indicate overall progression in males (blue) and females (pink) using the latest submission. Both male (blue squares) and female participants (pink circles) showed a rapid decline in their overall mobility with increasing age and disease duration.

#### Cognitive Function

Next, we assessed the cognitive function of the seventy participants who completed the Neuro-QoL cognitive function survey. PROM questions to assess cognitive function include ability to perform specific tasks such as reading and following complex instructions, concentrating, and paying attention. Similar to the technique used in the calculation of functional ability, we calculated the total raw scores and then created a scatter plot to show the correlation with age in years (Figure 5). Both males (N=33) and females (N=37) displayed an overall gradual decline in cognitive function, however females showed a slightly more rapid decline (slope=-0.18, R^2^=0.06) compared to males (slope=-0.06, R^2^=0.01) (Figure 5a). Overall, there is no correlation between cognitive function and disease duration for every year of age, regardless of sex (R^2^=0.01) (Figure 5b).

**Figure 5.**
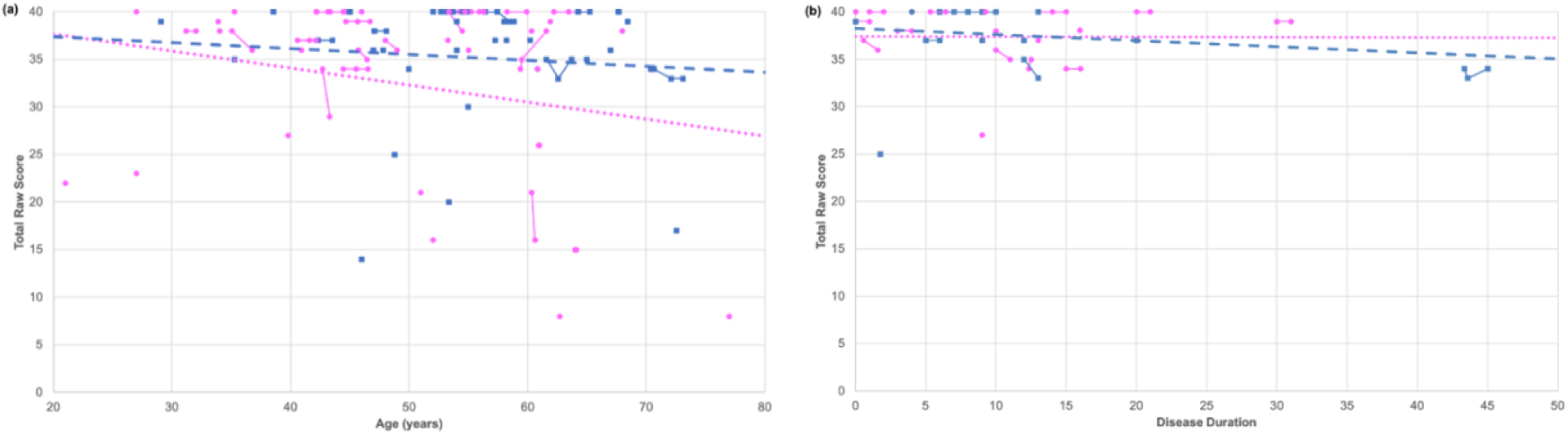
Cognitive Function Progression.

A scatter plot was created to illustrate the correlations between **(a)** age and **(b)** disease duration relative to the total reported raw score for cognitive function. The solid lines indicate progression within a singular participant over time for data with multiple submissions whereas the dashed lines indicate overall progression in males (blue) and females (pink) using the latest submitted value. Both male (blue squares) and female participants (pink circles) showed a decline in their overall cognitive abilities with increasing age whereas little to no decline with increasing disease duration.

#### Pain Level

Based on the overall pain scale, 13.0% of participants reported no pain, 38.2% reported mild pain (pain score1-3), 35.3% reported moderate pain (pain score 4-6), and 13.2% reported severe pain (pain score 7 or higher). Furthermore, 80.3% of symptomatic participants reported pain sometimes, often, or always interferes with their enjoyment of life (Figure S2). Using the latest participant data entries, we created a bar graph comparing the frequency of reported pain levels between male (N=34) and female (N=38) participants (Figure 6). There was a wide distribution in pain levels among participants, with 2 being the most frequent score for females (22.9%) and 3 being the most frequent score for males (24.2%), according to the latest submission. Interestingly, females showed a gradual decrease in pain (slope=-0.03, R^2^=0.03), whereas males showed an increase in pain with increasing age (slope=0.08, R^2^=0.13) (Figure 7a). Most male and female participants with multiple entries reported no change in pain, while some reported an increase in pain.

**Figure 6.**
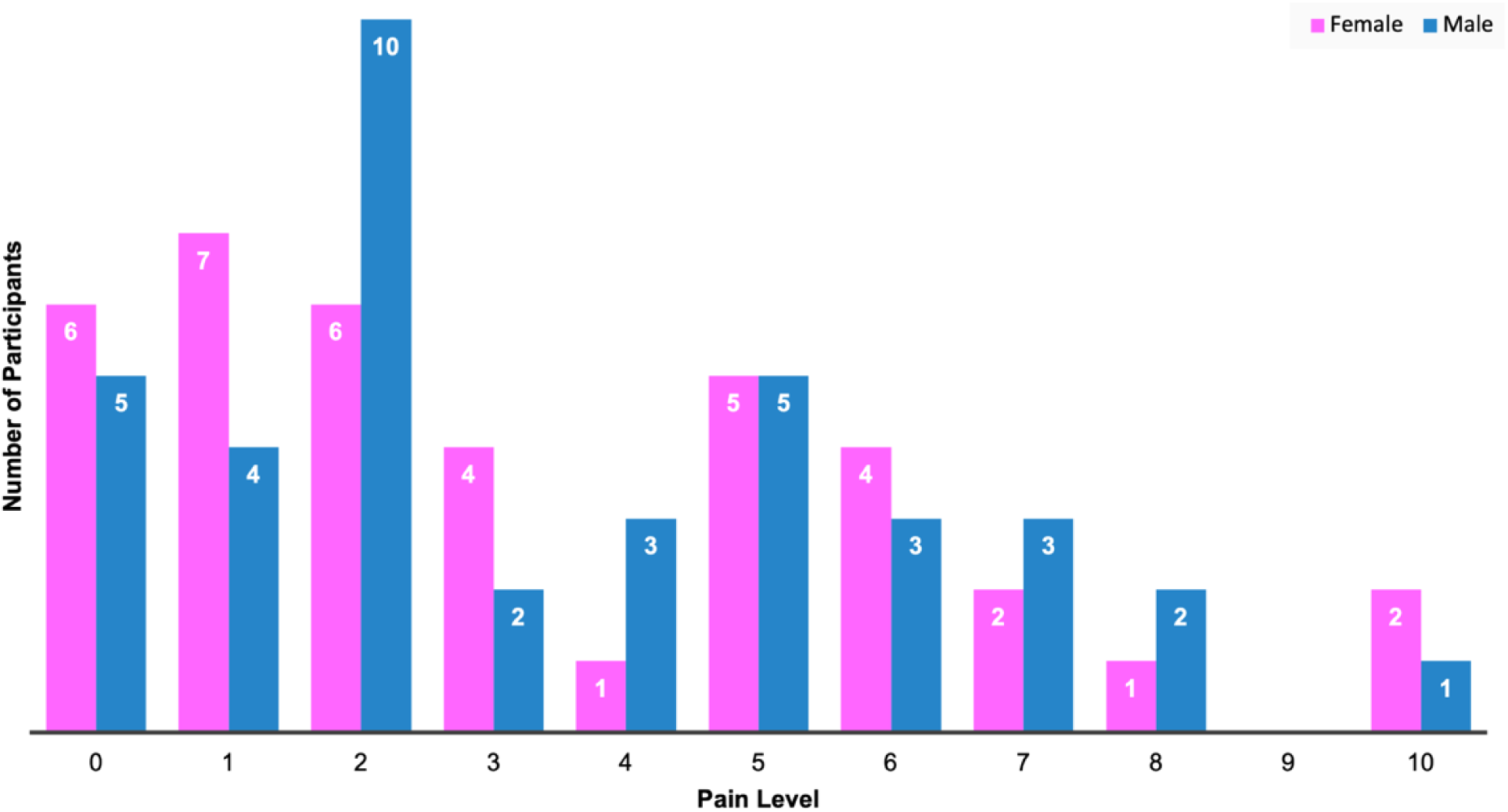
Comparison of Pain Levels in Males and Females. The bar graph shows the distribution of pain scores amongst males (blue) and females (pink). Pain levels of participants were recorded over the course of up to three years with the latest score being used for analysis. the pain scores ranged from 0-10, a value of 0 indicating no pain while a value of 10 indicating the worst pain imaginable.

**Figure 7.**
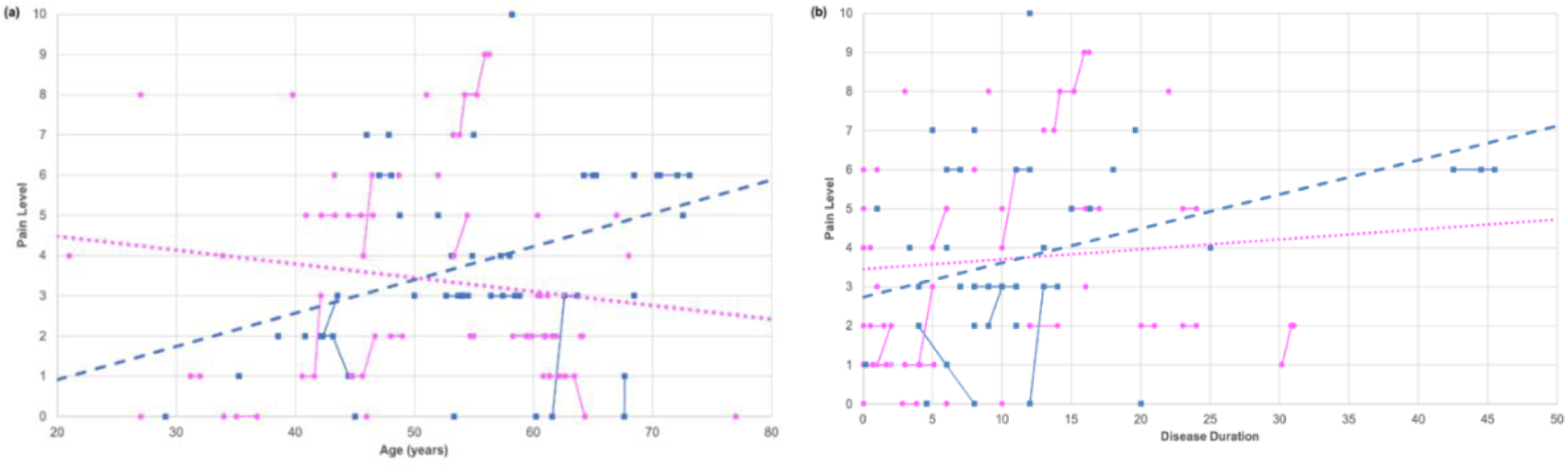
Pain Level Progression. A scatter plot was generated to illustrate the correlations between age **(a)** and disease duration **(b)** relative to the total reported raw score for pain. The solid lines indicate progression within a singular participant over time for data with multiple submissions whereas the dashed lines indicate overall progression in males (blue) and females (pink) using the latest submission. Male participants (blue squares) showed an incline in their overall pain levels with increasing age and disease duration whereas female participants (pink circles) showed a decline in pain levels with increasing age but an increase in pain levels as the disease progressed.

### Quality of Life

#### Quality of Life in Symptomatic Participants

Seventy-five participants completed the five QOL survey questions in the VCP CoRDS Registry (9 pre-symptomatic, 66 symptomatic) (Figure S2). Among symptomatic participants, 88% (57/66) of symptomatic participants reported that their health often or always limits doing vigorous activities, 76% (51/66) reported that they often or always felt tired, 55% (36/66) reported fair or poor general health, 45% (29/66) reported that pain always or often interfered with their enjoyment of life, and 29% (19/66) reported being always or often depressed. The responses of pre-symptomatic patients are included in the graph for reference, but due to the low numbers, a comparison was not calculated.

#### Quality of Life, Disease Progression, and Age Correlations

Moderate positive correlations were found between vigorous activities and upper (0.57) and lower (0.63) extremity function, suggesting that musculoskeletal health and functional ability across different body areas for patients with VCP disease has a negative impact on the intensity of physical exercise. Furthermore, the moderate positive correlation between pain interference and fatigue (0.63) implies that as pain interference increases, there is a negative impact on the enjoyment of life for individuals experiencing these symptoms. Depression and fatigue (0.46), and pain interference and depression (0.41) also had positive correlation. This suggests that those who often felt that pain interfered with their enjoyment of life also reported feeling depressed and those who reported often having feelings of depression also reported greater feelings of tiredness. This connected relationship emphasizes the concept that perceptions of pain interference, feelings of depression, and tiredness are closely intertwined, highlighting the complexity of these emotional and physical states VCP patients experience. Additionally, several moderate positive correlations were found between overall general health and other factors: lower extremity function (0.63), upper extremity function (0.46), fatigue (0.46), and vigorous activities (0.46). Moderate negative correlations were found between pain level and fatigue (−0.57), pain level and pain interference (−0.56), lower extremity function and age (−0.49), and vigorous activities and age (−0.41) while all other correlations were weak. This analysis provided a sophisticated understanding of the complex relationships among the factors under investigation. These findings emphasize the significance of considering multiple perspectives of VCP patients to improve affected individuals’ overall well-being and quality of life.

When comparing age, disease progression and QOL factors, a strong correlation was discovered between lower extremity function and upper extremity function (0.85). Additionally, moderate correlations were found between lower extremity function and vigorous activities (0.63). These findings are consistent with the known progression of muscle disease in both the upper and lower skeletal muscles. When analyzing patients’ determination of their overall general health, lower extremity function had the strongest correlation (0.63), with upper extremity function, vigorous activities and fatigue also revealing a moderate correlation to overall general health (0.46 respectively). Loss of muscle function has a direct effect on a person’s ability to complete daily activities and participate in vigorous activities, so these findings are not surprising. Our data revealed moderate correlations between age and lower extremity function as well as vigorous activities, while pain, general health, cognition function, and fatigue showed weak correlations with age.

#### Heat Map depicting Quality of Life correlations between key health factors

We studied correlations between pain level, cognitive function, lower extremity, upper extremity, general health, vigorous activities, pain interference, fatigue, depression factors that were evaluated in the questionnaire. The larger and darker the circle around the correlation value, the stronger the correlation between the two variables (Figure 8).

**Figure 8.**
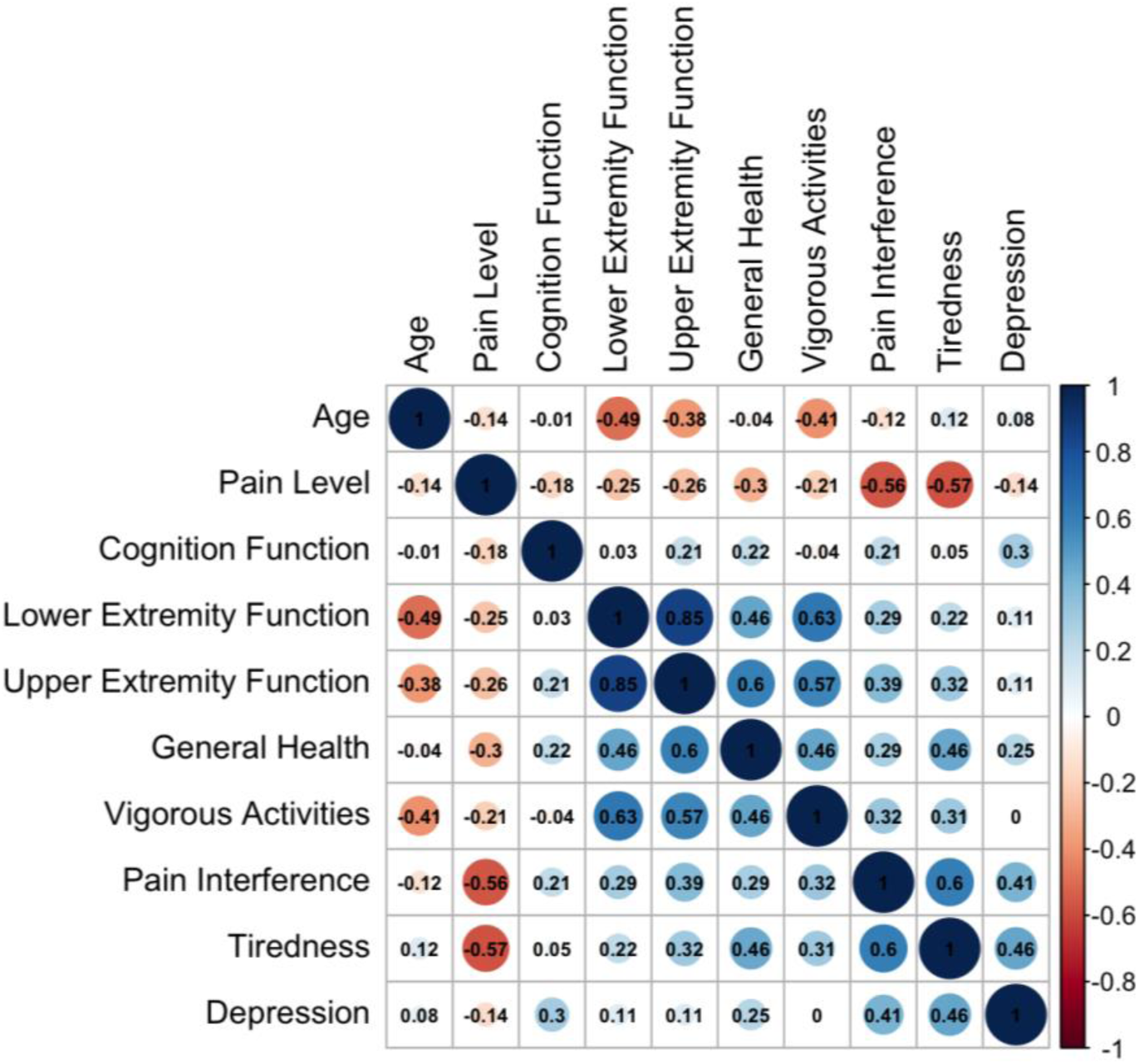
Quality of Life. This is a representation of a heat map signifying the correlation between the first five variables, which were measured during the study. General Health, Vigorous Activities, Pain Interference, Tiredness, Depression were factors that were evaluated from our questionnaire. The scale on the right represents the direction and intensity of the correlation. The larger and darker the circle around the correlation value, the stronger the correlation between the two variables.

A patient’s pain level displayed a moderate negative correlation to both fatigue (−0.57) and how often pain interfered with their enjoyment of life (−0.56). In other words, as pain level increased, a patient was more likely to report being tired and having pain interfere with their enjoyment of life. Strong evidence of a relationship between pain and tiredness, and between pain interference and depression. Interestingly, fatigue and pain interference have the same correlation to pain level suggesting the significance of both pain and fatigue in the enjoyment of life. Moreover, a significant percentage of VCP patients reported experiencing fatigue at high levels, with 76% (51/66) reporting that they often or always felt tired and 88% (57/66) of symptomatic participants reporting that their health often or always limits doing vigorous activities. A correlation analysis between fatigue, pain inference, and depression further revealed a moderate correlation between all three of these factors [pain inference and fatigue (0.63), fatigue and depression (0.46), and pain inference and depression (0.41)].

#### Overall Summary of Findings

Among the 79 participants, myopathy was the most prevalent phenotype (84.0%), followed by PDB (43.4%), FTD (12.9%), ALS (4.8%), and Parkinson’s disease (1.6%). Regarding the disease progression studies, participants revealed a significant decline of 0.6% in upper extremity function, 1.2% in lower extremity function, and 0.3% in cognitive function per year of age. Participants also revealed a 1.6% decline in upper and lower extremity function and a 0.1% decline in cognitive function per year of disease duration. This study highlights the high frequency of pain and the impact of fatigue on the quality-of-life aspects, such as depression and enjoyment of life in this patient population. Interestingly, the strongest correlation concerning patient reported general health was not age but lower extremity function. The *c. 464G>A, p.R155H* VCP variant was reported as the most prevalent variant group with the highest number of phenotypic characteristics. No obvious genotype-phenotype correlations for VCP disease phenotypes were noted.

## Discussion

Studying the natural history of a disease is important because it provides critical information on how the disease affects a patient’s daily life, understanding disease progression, which can thus be useful in creating preventative interventions, and permits studying the effects of therapeutic interventions. The inclusion of real-world evidence in the 21^st^ Century Cures Act and the use of PROMs in the FDA guidance in drug development elevates the need to evaluate natural history from the patient’s perspective early in the drug discovery process [17, 18].

### Influence of Genotype on Phenotype

Valosin-containing protein (*VCP)* variants can cause adult-onset proximal myopathy, which leads to upper and lower extremity weakness, axial myopathy, scapular winging, PDB and frontotemporal dementia (IBMPFD) [4, 19–21]. The most common phenotype was myopathy with the mean age of onset of 43.1 years as previously reported [22], followed by PDB with an average age of onset of 44.8 years. VCP has several functions: it is involved in cell division, repairing DNA damage, membrane fusion, and reassembling structures in cells post-division. It is also involved as an ATPase in autophagy and cell degradation, including ubiquitin-related cellular processes [5]. The VCP enzyme consists of the N domain and D1 and D2 domains, which are the ATPase modules. The D1 and D2 domains of VCP form a homohexamer in a head-to-tail ring structure, which allows the N domain to undergo conformational changes when ubiquitin-binding occurs. Missense variants in the *VCP* gene that cause IBMPFD can disrupt the double-psi barrel (R95G), four-stranded beta-barrel (R155C, R155H, R155P), or the flexible linker (R191Q). Most of the variants are in the N-terminal domain, which has the function of binding cofactors and ubiquitylated proteins [4]. We reported 13 unique variants in this cohort. The R155 codon is a mutational hotspot reported in 46% of the VCP variants reported in our study; the most prevalent is R155H, followed by R155C, as reported previously [4, 21]. Based on our genotype-phenotype comparisons study, we did not observe any significant correlations.

### Disease Progression

A notable decline in mobility was detected in participants with VCP disease, exhibiting a negative correlation with both advancing age and prolonged disease duration. The presence of comorbidities such as myopathy, PDB, ALS, and Parkinson’s disease further exacerbates the decline in mobility seen in those with VCP disease. The variability in decline of mobility due to VCP myopathy aligns with the documented progression pattern in limb-girdle muscular dystrophy (LGMD), which could be characterized by a heterogeneous onset of muscle weakness affecting the pelvic and shoulder girdles, typically manifesting in adolescence or early adulthood. The progression of LGMD varies widely, with some patients experiencing rapid deterioration while others show a slower decline [23, 24]. Variability in progression rate is similarly observed in ALS, particularly among patients with SOD1 mutations. For instance, individuals harboring the SOD1 A4V mutation often experience a more aggressive disease trajectory, with a median survival of approximately 1.2 years, in contrast to the 6.8 years observed for other SOD1 mutations [25]. Studies emphasize the importance of early clinical markers, including changes in functional scores and respiratory function, to elucidate disease trajectories and inform therapeutic strategies. These insights highlight the necessity of an integrated approach to the clinical management of VCP disease, addressing both the primary disorder and its associated co-morbidities to enhance patient outcomes and quality of life.

Overall, both male and female participants displayed slight cognitive impairments with age. This is consistent with previous studies of families with patients who displayed cognitive impairments at a mean age of 56 years [8]. Following a detailed investigation of the link between aging and cognitive performance, our findings demonstrated a steady deterioration in cognitive function with increasing age. Previous research confirms this by concluding a negative connection between age and cognitive performance across a variety of activities, including memory, processing speed, and executive functioning [26]. The research goes into neuroimaging studies, which highlight structural and functional changes in the aging brain, notably in memory and information processing areas [26]. According to another study, the aging brain experiences complex interactions and changes that affect many cognitive processes other than memory and information processing. Altogether, these investigations confirm a similar trend: an individual’s cognitive capacity tends to deteriorate with age. In addition to identifying potential treatment and preventive methods to reduce age-related cognitive loss, it is critical to underline the importance of multidisciplinary research in understanding the complex nature of cognitive aging [27].

### Pain Implications

In analyzing the reported pain levels, males reported more pain with the progression of the disease (slope = 0.08 versus -0.03, p<0.01) with advancing age. In other studies, a multitude of factors, including biological, psychosocial, sex difference, have been proven to influence an individual’s pain, and understanding these factors can serve as a vital foundation for individualized pain treatment plan [28]. Pain is a common and underappreciated manifestation of VCP disease and can result from comorbidities such as PDB, which typically affects the lumbar spine, hips, pelvis and skull [25]. It is important to investigate the presence of PDB to reduce the additional burden since it is readily treatable with bisphosphonates. Interestingly in this cohort, symptomatic VCP participants with PDB did not report a higher level of pain when compared to symptomatic VCP participants without PDB (average pain score of 3.2/10, 4.1/10, respectively). These results may suggest that patients may have undiagnosed PDB or that some patients with PDB may be receiving treatment to address their pain. We further analyzed similarities among patients who reported high levels of pain, and the presence of myopathy was the only common factor we discovered. Interestingly, a greater percentage of VCP participants without PDB reported a moderate to high level of pain (4 or higher) when compared to VCP participants with PDB (55% 26/45, 37% 6/16). These results suggest that PDB may not be the main source of pain and that other disease-specific causes of pain exist in VCP disease, such as muscle pain or neuropathy. Thus, these results indicate that the detection and treatment of an individual’s pain needs to be part of a VCP patient’s clinical care plan; future studies are needed to determine the causes of pain in VCP patients.

### Sex Considerations

Other research studies indicate that males may have a more noticeable perception of decline in physical activity as they are unable to reach their activity goals, and this perception of change may explain the steeper reported decline in lower extremity function reported in males (slope=-0.52, R^2^=0.34) compared to the females (slope=-0.41, R^2^=0.22). One particular study found a difference in motivating factors for adult physical activities amongst the sexes [22]. Another study found that while males were motivated by mastery of physical activity, females were motivated by the psychosocial wellbeing component of exercise [26]. Additionally, previous studies have demonstrated that males were less likely to report depression than females, despite more males completing suicide due to the gravity of their deteriorating mental health [27]. However, we cannot ignore the potential biological differences between males and females and the prevalence of depression. There is evidence that estrogen acting as a psycho-protective component in the body, and the conversion of testosterone to estrogen in males mediates a protective action throughout life. While estrogen levels fluctuate with the premenstrual cycle in females, testosterone has a more consistent level in males [27]. Cognitive function results from this cohort showed a slightly more rapid decline in females (slope=-0.1782, R^2^=0.06) compared to males (slope=-0.0622, R^2^=0.01), but a low incidence of dementia reported in this cohort may influence these findings.

### Comparing Quality of Life Parameter Correlations

In a thorough investigation conducted by Yamada et al., cross-sectional studies of the data unveiled significant correlations between measures of pain and fatigue/tiredness [29]. Although the study’s main focus was on the relationship between pain and tiredness symptoms over time in people with musculoskeletal disorders, it is crucial to interpret these results in the context of the larger body of research on the relationship between pain and fatigue. Strong evidence of a relationship between pain and tiredness was found in our dataset; this relationship graphically displayed in a heatmap. This numerical insight not only underscores the significance of our study’s specific analysis but also contributes valuable quantitative evidence to the broader body of research affirming the intricate relationship between pain and fatigue.

According to Linton et al.’s analysis, there is a clear relationship between pain and depression, which calls for a complex understanding of their interaction within the therapeutic framework [30]. Within our dataset there was a significant relationship between pain interference and depression. These results highlight the necessity of further research efforts to thoroughly investigate this association, clarifying the importance for improving therapeutic approaches. Fatigue or exhaustion is a widespread issue in both community and medical settings, and there is a strong association with depression. The correlation between enhanced work performance and improved energy underscores the importance of addressing fatigue in the context of depression. In our study, an analysis of depression and tiredness/fatigue revealed a significant association meriting the need for further investigation [31].

Paterson et al.’s study of generalized physical activity and functional limitations demonstrated that regular physical activity is associated with a reduced risk of functional limitations, particularly in the older age group [32]. In a preclinical study of VCP disease, exercise training in mice with the *Arg155His* Vcp variant found that there was an improvement with uphill exercise [33]. Korb et al.’s recommendations of the standard of care for patients with VCP disease acknowledges that exercise has a positive effect on individuals with FTD [34]. A recent study on resistance respiratory exercise in subjects with VCP myopathy showed that the maximum inspiratory pressure increased significantly over the 32 week course of the resistance training compared to the baseline [35]. The effects of generalized exercise training however have not been studied and is a significant unmet need because of the absence of clear guidelines on exercise training in the VCP myopathy patient population.

### Patient Perspective and Management of Care

Quality of life is imperative to navigating a patient’s health. Treatment options are catered to maximizing a patient’s quality of life, and therefore understanding patient perspectives is integral to optimizing patient care [36]. In chronic diseases of neurological or musculoskeletal disabilities, it has been demonstrated that psychological impacts may lead to a decline in the QOL of a patient [37–38]. Additionally, chronic illnesses like VCP disease may increase the prevalence of depression. Routine management of chronic disease warrants depression screenings and management of pain as patients progress through life with their diagnosis [38]. To gain a better understanding of patient perspectives in VCP disease, we analyzed the responses to better understand how fatigue, pain, and depression directly impacted their daily life. A patient’s pain level displayed a moderate negative correlation to both fatigue (−0.57) and how often pain interfered with their enjoyment of life (−0.56). In other words, as pain level increased, a patient was more likely to report being tired and having pain interfere with their enjoyment of life. Interestingly, fatigue and pain interference have the same correlation to pain level suggesting the significance of both pain and fatigue in the enjoyment of life. Moreover, a significant percentage of VCP patients reported experiencing fatigue at high levels, with 76% (51/66) reporting that they often or always felt tired and 88% (57/66) of symptomatic participants reporting that their health often or always limits doing vigorous activities. A correlation analysis between fatigue, pain inference, and depression further revealed a moderate correlation between all three of these factors [pain inference and fatigue (0.63), fatigue and depression (0.46), and pain inference and depression (0.41)], suggesting that these symptoms may be interrelated and should be monitored throughout the life of a VCP patient. The inclusion of a mental health section in the VCP care guidelines further demonstrates the importance of including psychological health in a VCP patient’s overall care plan [34].

## Limitations and Future Directions

Understanding the mechanisms of complex traits through genotype-phenotype comparisons can be limiting because there can be multiple genetic and environmental factors that can contribute to the differences. Understanding interactions between modifier genes and variants on the gene-of-interest, epigenetics, and environmental factors that lead to certain phenotypes are factors that should be taken into consideration [21]. The phenotypic diversity of VCP disease is not necessarily due to the specific types of VCP variants [8]. Patients with the same *VCP* variant who are in the same family can present with different phenotypes and clinical manifestations. Other environmental or genetic modifiers that have yet to be investigated [24]. Prior studies have shown a correlation of the apolipoprotein-E (*APOE4*) allele as a potential VCP modifier gene [39]. VCP patients with one or more *APOE4* alleles had dementia as a phenotypic characteristic. Based on these studies, modifier genes and potential interactions with VCP such as *APOE4, NF1*, among others should be accounted for in future studies to better understand their effects on phenotype along with VCP variants [39]. Our group has also indicated that the R159C variant is associated with a lower incidence of PDB [16].

While the PROMs are very useful in understanding a patient’s perspective of disease and monitoring their longitudinal progression, there are limitations to this data. Because PROMs are subjective, there are many parameters that can influence the subject’s response. Depending on how patients have been feeling, even days leading up to the questionnaire, can influence their general perspective of how they answer the survey questions. Moreover, there can be variation in assessment on a day-to-day basis since the time period for this questionnaire is 7 days. In addition, every subject’s idea of items such as “vigorous activities”, “depression”, “general health”, and “pain” are subjective and based on their perception. In addition, participants have subjective levels of pain tolerance at baseline. Moreover, the progression of the disease does not have the same initial baseline for the participants, as the disease duration is unique for each participant. Therefore, progression is relative to the patient’s disease progression. For a more complete disease understanding and natural history, PROMS should be complemented with clinical examination, diagnostic imaging, and clinical diagnosis.

Analyzing the patient-reported progression of VCP disease can provide great insight into its clinical manifestations; however, more longitudinal data is needed. While we were able to observe the overall trend in cognitive function, mobility, and pain in this study, only 17 participants reported unique results two or more times. most participants only reporting one data entry. This limited our ability to report on an individual’s reported change over time. Retrieving additional data entries from existing participants will provide a better understanding of the disease progression. In addition, including more pre-symptomatic participants with a VCP disease variant will allow the monitoring of disease progression longitudinally. Another limitation is that participants did not submit answers to all survey questions due to the protocol management by the internal review board at Sanford Research. This protocol reviewed by the review board at Sanford Research allows subjects the option to omit questions which they do not feel comfortable answering. While this protocol may eliminate erroneous answers from patients and increase overall registry enrollment, not having all answers completed leads to challenges in conducting correlation analysis, including having to exclude some participants from calculations entirely.

Increasing the sample size of the cohort to include statistically significant data in VCP genotype-phenotype comparisons can help to analyze the progression in participants with other comorbidities such as ALS, PDB, and peripheral neuropathy. This could provide better insight as to how other conditions impact the severity of QOL symptoms amongst participants with VCP disease. Moreover, collecting data for a longer period of time will allow for increased data consistency and the monitoring of the long-term progression of VCP phenotypic manifestations. As more responses are collected over time in the VCP CoRDS Registry, further data analysis can be conducted, allowing for a longer span of time to understand the correlation of how QOL changes over time with disease progression.

## Conclusion

Patient registries are valuable tools in improving clinical care and setting future research direction by including the patient voice, capturing natural history data, and elevating the QOL of patients. The VCP CoRDS Registry was found to be a valuable tool for monitoring the quality of life in patients with VCP disease. The results of this study could be a useful tool in monitoring the natural history progression of upper extremity function, lower extremity function, and cognitive function in the VCP patient population. This study indicates that patient-driven registries such as CoRDS can be used to monitor disease progression in rare diseases on a global scale.

## Declarations

### Ethics Approval and Consent to Participate

Written informed consent was obtained from all individuals. This study was approved by the University of California Irvine Institutional Review Board (IRB) (#2007-5832).

### Consent for Publication

Written informed consent was obtained from all the subjects included in the study. In the case of deceased patients, they provided their consents prior to their demise.

### Authors’ Contributions

Virginia Kimonis, Allison Peck: Conceptualization, methodology, resources, supervision, validation, project administration, writing and funding acquisition.

Eiman Abdoalsadig: Investigation, writing - original draft, visualization, and validation. Merwa Hamid: Investigation, writing - original draft, visualization, and validation.

Leepakshi Johar: Investigation, writing - original draft, and visualization. All authors read and approved the final manuscript.

### Availability of Data and Materials

The datasets generated during and/or analyzed during the current study are available from the corresponding author on reasonable request.

## Data Availability

All data produced in the present study are available upon reasonable request to the corresponding author

## Acknowledgments

We thank the participants for their contribution to the study. Our gratitude extends to the Cure VCP Disease, Inc., Alyssa Mendel, and the Coordination of Rare Diseases at Sanford (CoRDS) Research Institute, Sioux Falls, S. Dakota for their support with the acquisition and management of registry data. We are grateful to Regina Im, Vu Luu, Mabel Tang, and Ryan Mahoney for their contribution to this work.

---

**Supplemental Table 1:**
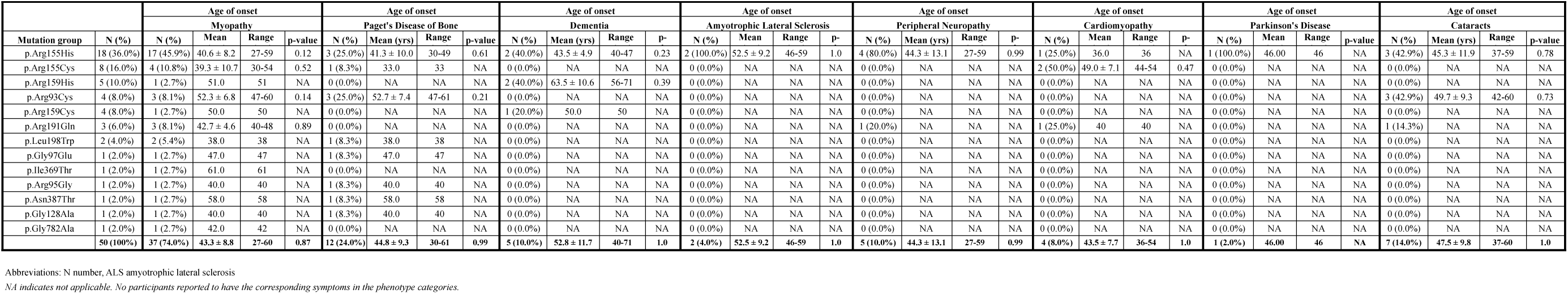
Genotype—Phenotype Comparison Data of Clinical Features 238 Symptomatic Patients in Different Variant Groups

